# In-silico analysis of differentially expressed genes and their regulating microRNA involved in lymph node metastasis in invasive breast carcinoma

**DOI:** 10.1101/2020.11.25.20235259

**Authors:** Anupama Modi, Purvi Purohit, Ashita Gadwal, Shweta Ukey, Dipayan Roy, Sujoy Fernandes, Mithu Banerjee

**Author notes:** Corresponding author Dr. Purvi Purohit, Additional Professor, Department of Biochemistry, AIIMS, Jodhpur, Rajasthan- 342005. Funding: None.

## Abstract

**Introduction:** Axillary nodal metastasis is related to poor prognosis in breast cancer (BC). The metastatic progression in BC is related to molecular signatures. The currently popular methods to evaluate nodal status may give false negatives or give rise to secondary complications. In this study, key candidate genes in BC lymph node metastasis have been identified from publicly available microarray datasets and their roles in BC have been explored through survival analysis and target prediction.

**Methods:** Gene Expression Omnibus datasets have been analyzed for differentially expressed genes (DEGs) in lymph node-positive BC patients compared to nodal-negative and healthy tissues. The functional enrichment analysis was done in database for annotation, visualization and integrated discovery (DAVID). Protein-protein interaction (PPI) network was constructed in Search Tool for the Retrieval of Interacting Genes and proteins (STRING) and visualized on Cytoscape. The candidate hub genes were identified and their expression analyzed for overall survival (OS) in Gene Expression Profiling Interactive Analysis (GEPIA). The target miRNA and transcription factors were analyzed through miRNet.

**Results:** A total of 102 overlapping DEGs were found. Gene Ontology revealed eleven, seventeen, and three significant terms for cellular component, biological process, and molecular function respectively. Six candidate genes, DSC3, KRT5, KRT6B, KRT17, KRT81, and SERPINB5 were significantly associated with nodal metastasis and OS in BC patients. A total of 83 targeting miRNA were identified through miRNet and hsa-miR-155-5p was found to be the most significant miRNA which was targeting five out of six hub genes.

**Conclusion:** In-silico survival and expression analyses revealed six candidate genes and 83 miRNAs, which may be potential diagnostic markers and therapeutic targets in BC patients and miR-155-5p shows promise as it targeted five important hub genes related to lymph-node metastasis.

## 1. Introduction

According to the latest 2018 report of GLOBOCAN, breast cancer (BC) is the most common cancer in females which accounts for 11.6% among all cancer types. BC is commonly invasive, and there are 626,679 deaths due to BC worldwide, making it the second major cause of cancer-related deaths [1]. The involvement of axillary lymph nodes leads to worse prognosis in BC. According to the American Joint Committee on Cancer (AJCC)/International Union Against Cancer (UICC) tumor (T)-node (N)- metastasis (M) classification, nodal disease is classified in three groups based on the number of axillary metastatic lymph nodes involved: N1, 1–3 metastatic lymph node(s), N2, 4–9 metastatic lymph nodes, and N3, 10 or more metastatic lymph nodes [2]. The prognosis worsens with the increase in the number of metastatic lymph nodes [3, 4]. Five-year survival rates differ dramatically between women with negative lymph nodes (>90%) compared to those with lymph node metastasis (<70%) [5].

Positive or negative lymph node metastatic status aid in patient staging, prognostic information, and patient management [6]. Bakkour et al. observed that axillary lymph node involvement is significantly associated with overall survival (OS) and disease-free survival (DFS) [4] Studies have also reported that a rise in the number of nodal involvement increases the rate of cancer recurrence [7, 8]. Although the involvement of axillary lymph node is the prognostic indicator of BC, the current method used to identify the nodal status, sentinel lymph node biopsy (SLNB), may give false-negative results, disrupt the lymphatic system and may lead to secondary complications. Therefore, there is a need for an appropriate method to identify the patients with and without lymph node metastasis, which could also reduce the chances of comorbidity related to surgical evaluation [9]. Identification of signature genes involved in lymph node metastasis of BC may be helpful in this process.

Differential gene expression signature can predict the tumor grade, subtypes and recurrence [9,10], which can predict the prognosis of diseases. Signature genes involved in local metastases in lymph node may not predict the outcome but it would have clinical and biological significance and improve our understanding of the process of metastasis [10]. Further, these genes could serve as therapeutic targets for the treatment of early-stage metastasis.

In this present study, we aimed to identify the crucial genes involved in lymph node metastasis in BC through publicly available microarray gene expression datasets available from the Gene Expression Omnibus (GEO). Through an in-silico approach, Kaplan-Meier survival analysis of hub genes was performed to identify the role of hub genes in the prognosis of BC. Further, for the hub genes which were significantly associated with poor prognosis of BC, their targeting miRNAs and transcription factors were identified.

## 2. Materials and Methods

### 2.1 Data collection

Gene expression profiles of axillary lymph node (N^-^) (GSE42568, GSE22093, GSE76275, GSE23988, and GSE36771) and non-axillary lymph node involving (N^+^) BC tissue, and healthy breast tissue (GSE42568) were downloaded from public database GEO [11-13]. These datasets were classified into three groups: 1) N- vs. control tissues (GSE42568) which included 45 N^-^ BC tissues and 17 control breast tissues, 2) N^+^ vs. control tissues (GSE42568) contained 59 N^+^ BC tissues and 17 control breast tissues, and 3) N^-^ vs. N^+^ (GSE42568, GSE22093, GSE76275, GSE23988, and GSE36771) including a total of 219 N^-^ and 250 N^+^ BC tissues, respectively (Table 1).

**Table 1:** Group-wise distribution of breast cancer tissue sample obtained from each dataset

| S.No. | GEO Dataset | No. of control<br>samples | No. of N<sup>+</sup><br>samples | No. of N<sup>-</sup><br>samples |
| --- | --- | --- | --- | --- |
| 1. | GSE42568 | 17 | 59 | 45 |
| 2. | GSE23988 |  | 40 | 21 |
| 3. | GSE22093 |  | 30 | 21 |
| 4. | GSE76275 |  | 76 | 74 |
| 5. | GSE36771 |  | 45 | 58 |
|  | Total | 17 | 250 | 219 |

### 2.2 Identification of differentially expressed genes (DEGs)

The interactive web tool GEO2R (https://www.ncbi.nlm.nih.gov/geo/geo2r/) was used to compare between groups of samples for differential expression analysis. The DEGs between N^-^ vs. Control, N^+^ vs. Control and N^-^ vs. N^+^ groups from GSE datasets were determined according to the cut-off value of |log_2_fold change (FC)| ≥0.58 and *P-*value <0.05. For visualization of DEGs, volcano plot was constructed for each dataset using R [14] (Figure 2a-g). Venn diagram was constructed using Venn diagram tool available at http://bioinformatics.psb.ugent.be/webtools/Venn/ for identifying the overlapping DEGs. Workflow for processing of data has been shown in figure 1.

**Figure 1:**
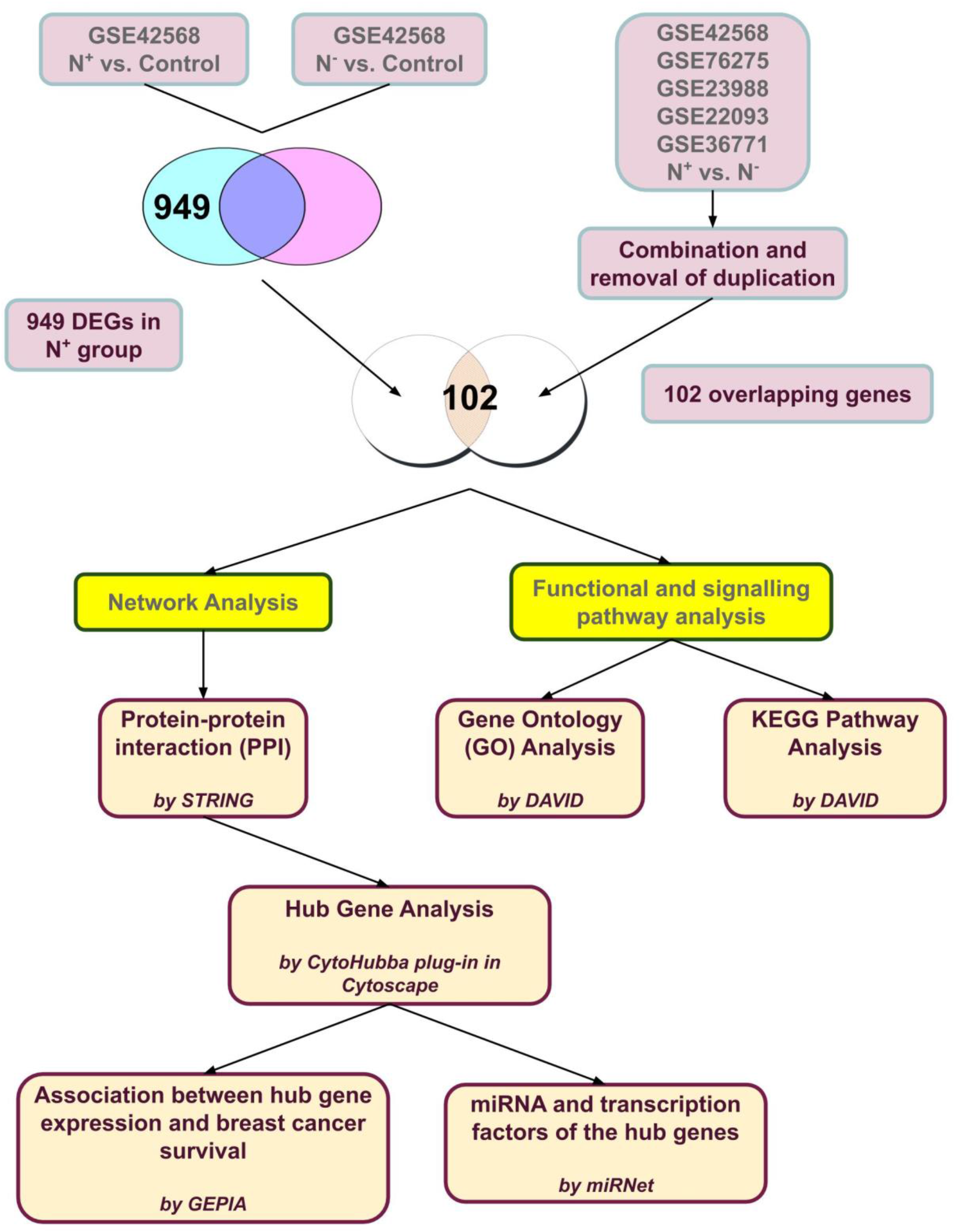
A schematic diagram of the workflow

**Figure 2a-g:**
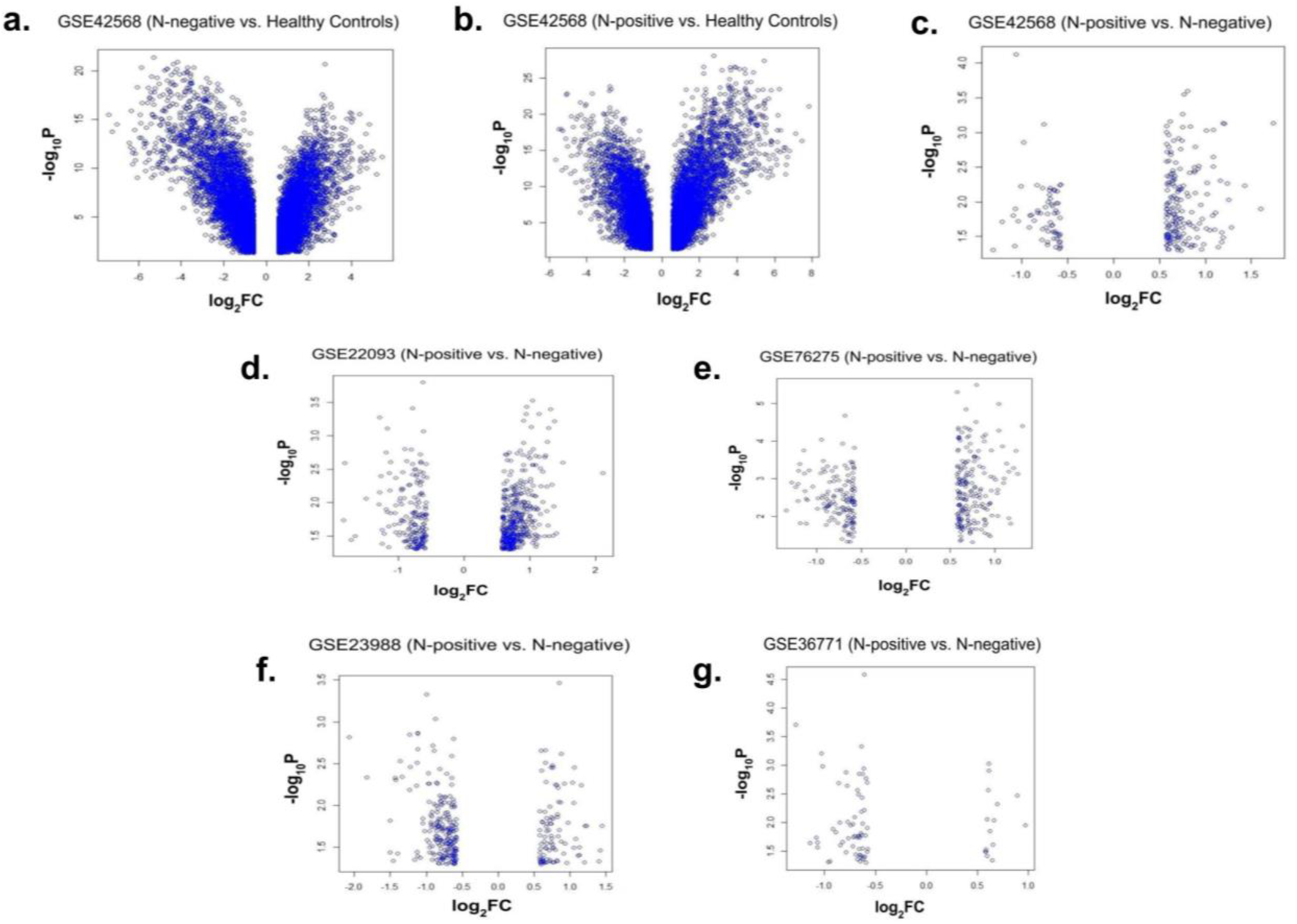
Volcano plots for the differentially expressed genes (DEGs) for each dataset

### 2.3 Functional Enrichment of DEGs

Gene ontology (GO) and Kyoto Encyclopedia of Genes and Genomes (KEGG) pathway functional enrichment were analyzed through the database for annotation, visualization and integrated discovery (DAVID; https://david.ncifcrf.gov/) [15]. *P-*value <0.05 was considered to be statistically significant. For the overlapping DEGs, GO enrichment analyses were classified into three functional groups: biological processes (BP), cellular components (CC), and molecular functions (MF).

### 2.4 PPI network construction

Protein-protein Interaction (PPI) network of overlapping 102 DEGs was constructed using the Search Tool for the Retrieval of Interacting Genes and proteins (STRING) database (https://string-db.org/) [16] which analyse the functional interaction between proteins. To explore the regulatory mechanisms, interactions with the confidence of a combined score >0.400 were retained and imported to Cytoscape (version 3.8.0) [17] for visualization of the PPI network.

### 2.5 Hub gene analysis

In Cytoscape, the CytoHubba plug-in, which uses 12 different methods and provides a user-friendly interface to analyze the topology of PPI networks to select the top 10 genes, was employed [18]. Those genes which were detected with at least three different methods were considered as the hub genes.

### 2.6 Expression and survival analysis of hub genes

Gene Expression Profiling Interactive Analysis (GEPIA) (http://gepia.cancer-pku.cn/) [19] is an interactive web server, which analyzes RNA-sequence expression data of 9,736 tumors and 8,587 normal samples from The Cancer Genome Atlas (TCGA) and Genotype-Tissue Expression (GTEx) projects. It can also conduct OS by log-rank test based on relative gene expression. The Cox proportional hazard ratio (HR) and the 95% confidence interval (95% CI) of the survival plot can also be obtained. In our study, we explored the OS of individual hub genes through the Kaplan–Meier plotter and tissue expression of hub genes in BC.

### 2.7 Hub gene targeting miRNA and transcription factor analysis

Hub gene targeting miRNAs were predicted by using miRNet 2.0 database (https://www.mirnet.ca/miRNet/home.xhtml), a miRNA-centric network visual analytics platform, and hub gene targeting transcription factors were analysed using miRNet at five different platforms (CHEA, ENCODE, JASPAR, REGNETWOTK and TRRUST). Further, hub gene-miRNA and hub gene-transcription factor interaction networks were visualized in Cytoscape.

## 3. Results

### 3.1 Identification of DEGs

For the chosen datasets, the number of DEGs were 7935 (GSE42568, N^-^ vs. control), 8298 (GSE42568, N^+^ vs. control), 221 (GSE42568, N^-^ vs. N^+^), 292 (GSE76275), 333 (GSE23988), and 551 (GSE22093), respectively. The Venn diagram for DEGs was constructed which showed 949 genes in the N^+^ group. To identify the genes differentially expressed in N^+^ vs. N^-^ group, from data sets (GSE42568, GSE22093, GSE76275, GSE23988 and GSE 36771) DEGs were downloaded. These DEGs were combined, and after removing the duplicate genes, there were 1287 DEGs. For further analysis, Venn diagram of 949 genes which were differentially expressed in N^+^ BC compared to controls, and 1287 DEGs of N^+^ vs. N^-^ group was constructed, which revealed that there were 102 overlapping DEGs based on the cut-off criteria of |log _2_FC| ≥0.58 and *P* value<0.05 (Figure 3).

**Figure 3:**
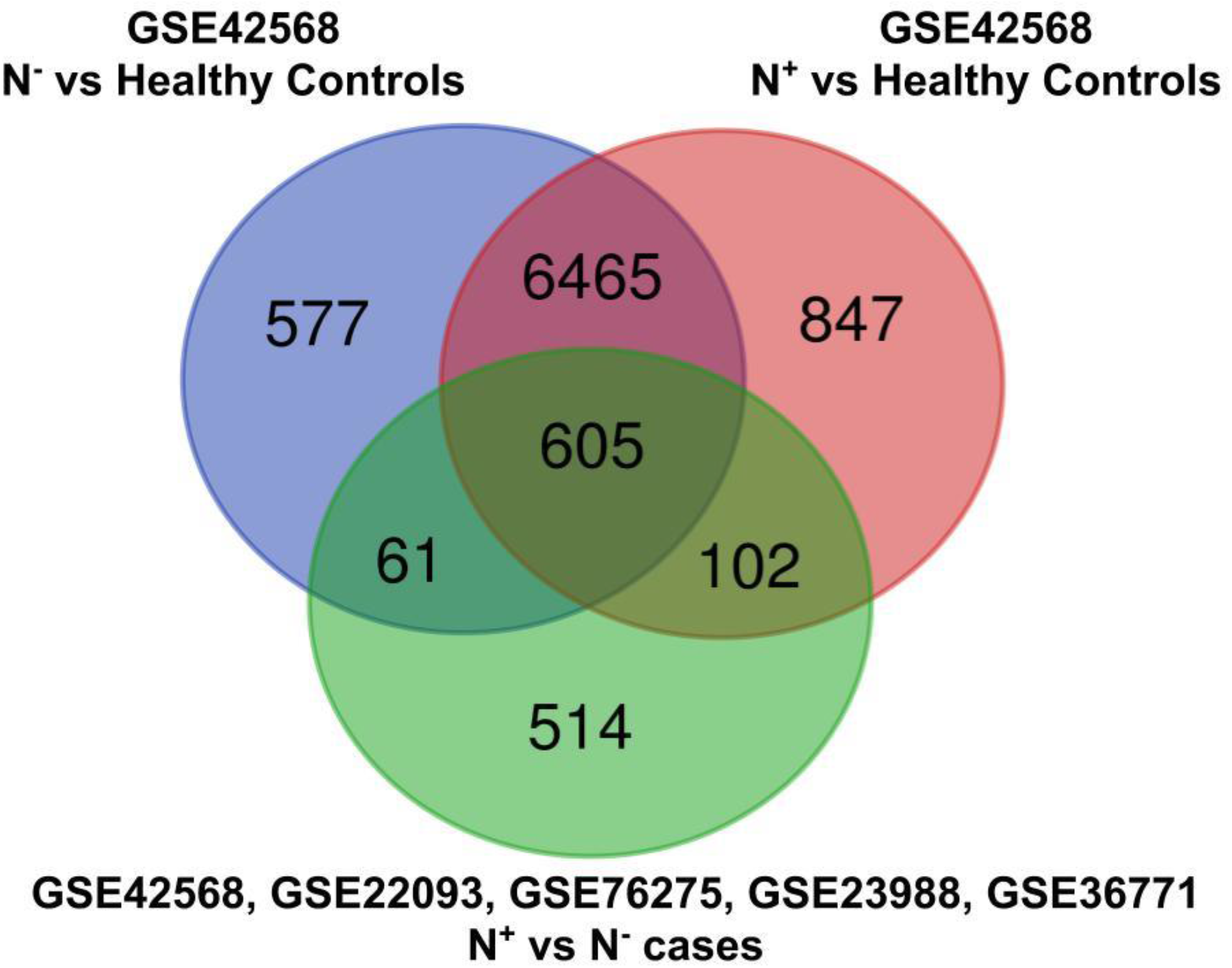
Venn diagram depicting the 102 overlapping DEGs for N^+^ BC patients

### 3.2 Functional enrichment analysis of DEGs

Three GO terms: CC, BP, and MF were analysed. KEGG pathways were analysed to identify the pathways related to the DEGs. In CC, BP, and MF, eleven, seventeen, and three GO terms, respectively, were found to be statistically significant. For CC, most of the genes were involved in Immunoglobin complex, circulating; Blood microparticles; External side of plasma membrane; Extracellular exosome; Extracellular space; whereas for BP, there was Positive regulation of B-cell activation; Phagocytosis, recognition; Phagocytosis, engulfment; B cell receptor signalling pathway; Complement activation, classical pathway. For MF, the significant terms were Immunoglobin receptor binding, Antigen binding, and Serine-type endopeptidase activity. Similarly, KEGG pathway enrichment analysis revealed that the DEGs were mainly involved in the p53 signalling pathway and Aldosterone synthesis and secretion (Figure 4a-d, Table 2-5).

**Table 2:**
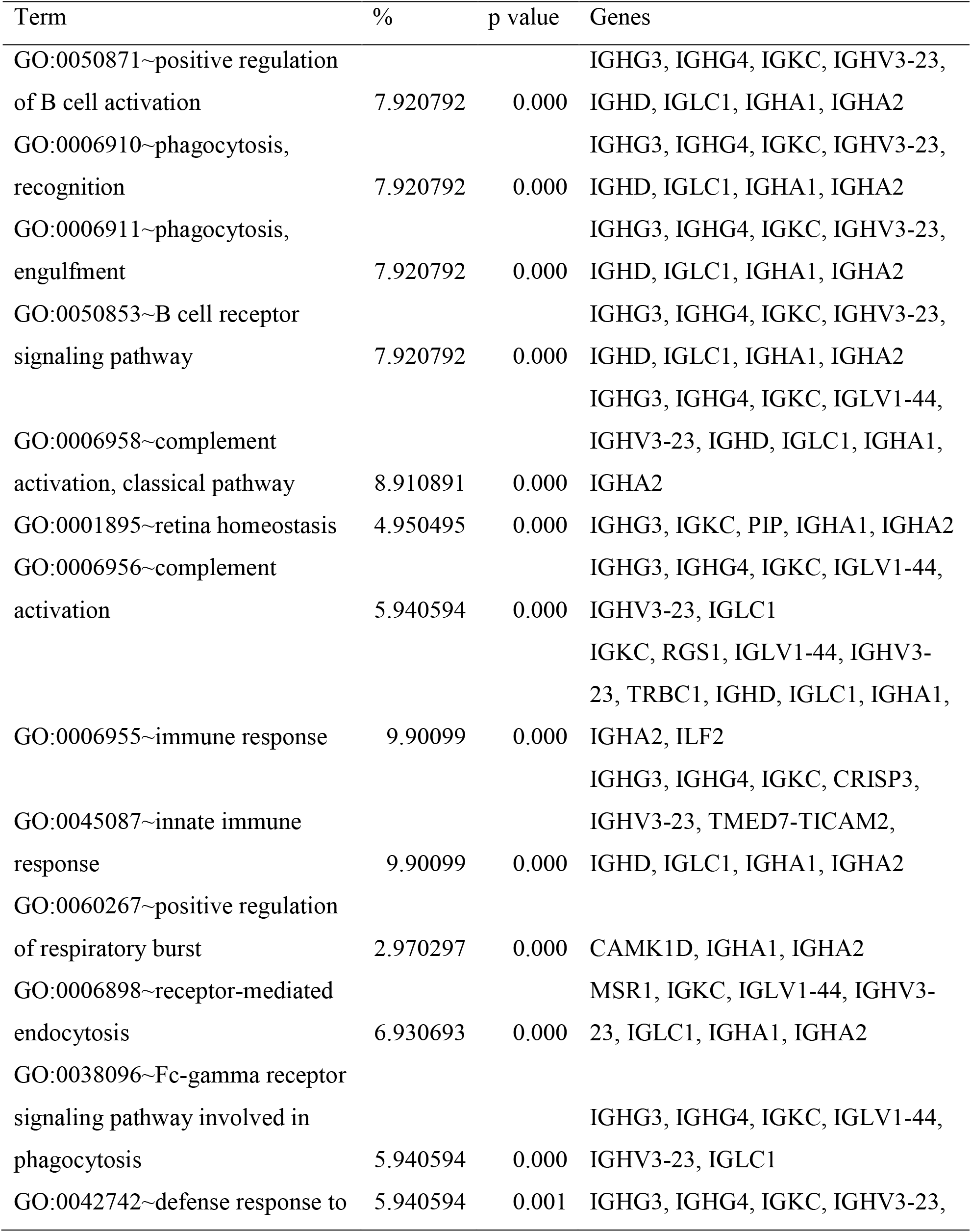
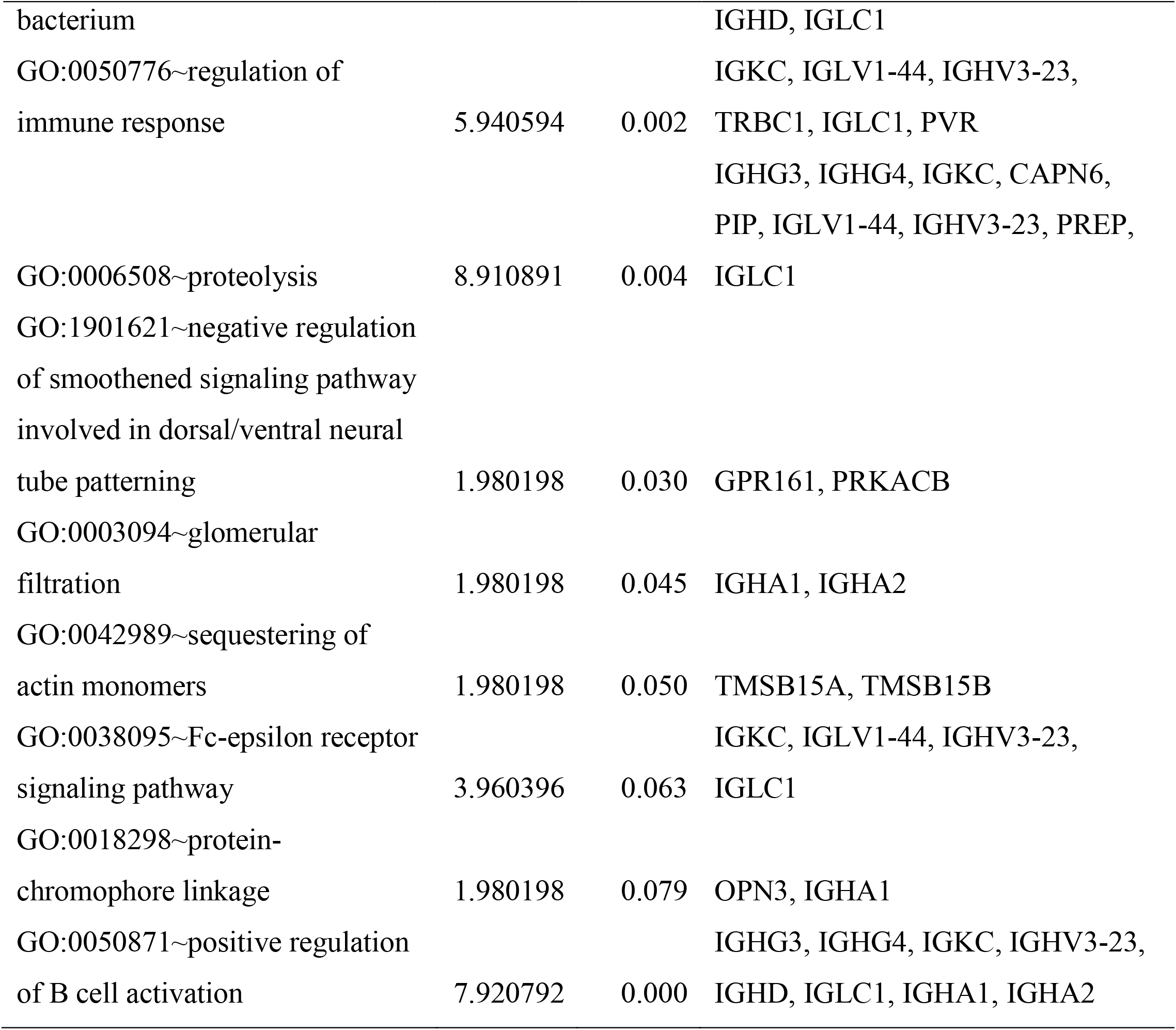
Gene Ontology analysis for the overlapping genes ∼ Biological Process

**Table 3:**
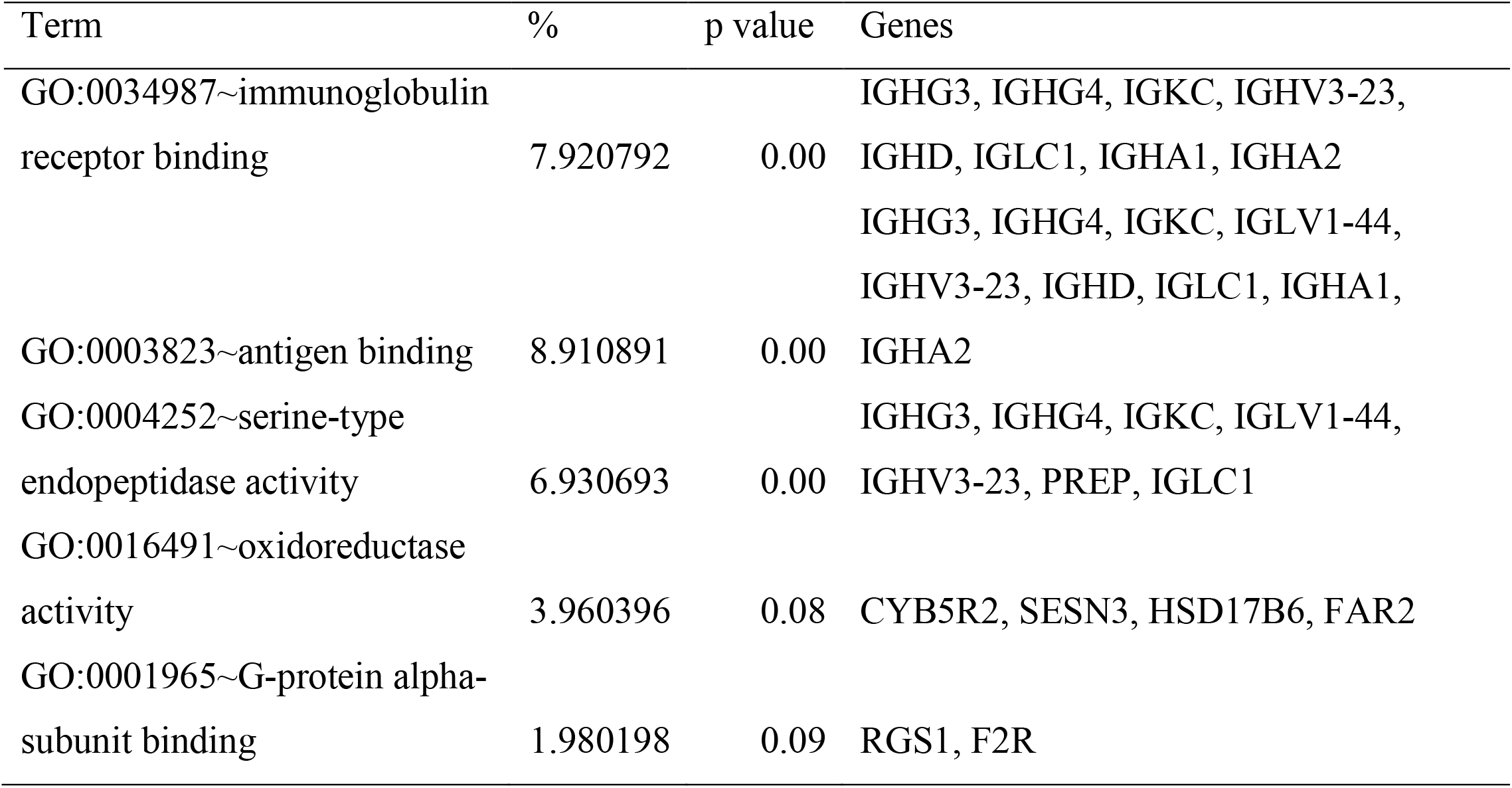
Gene Ontology analysis for the overlapping genes ∼ Molecular Function

**Table 4:**
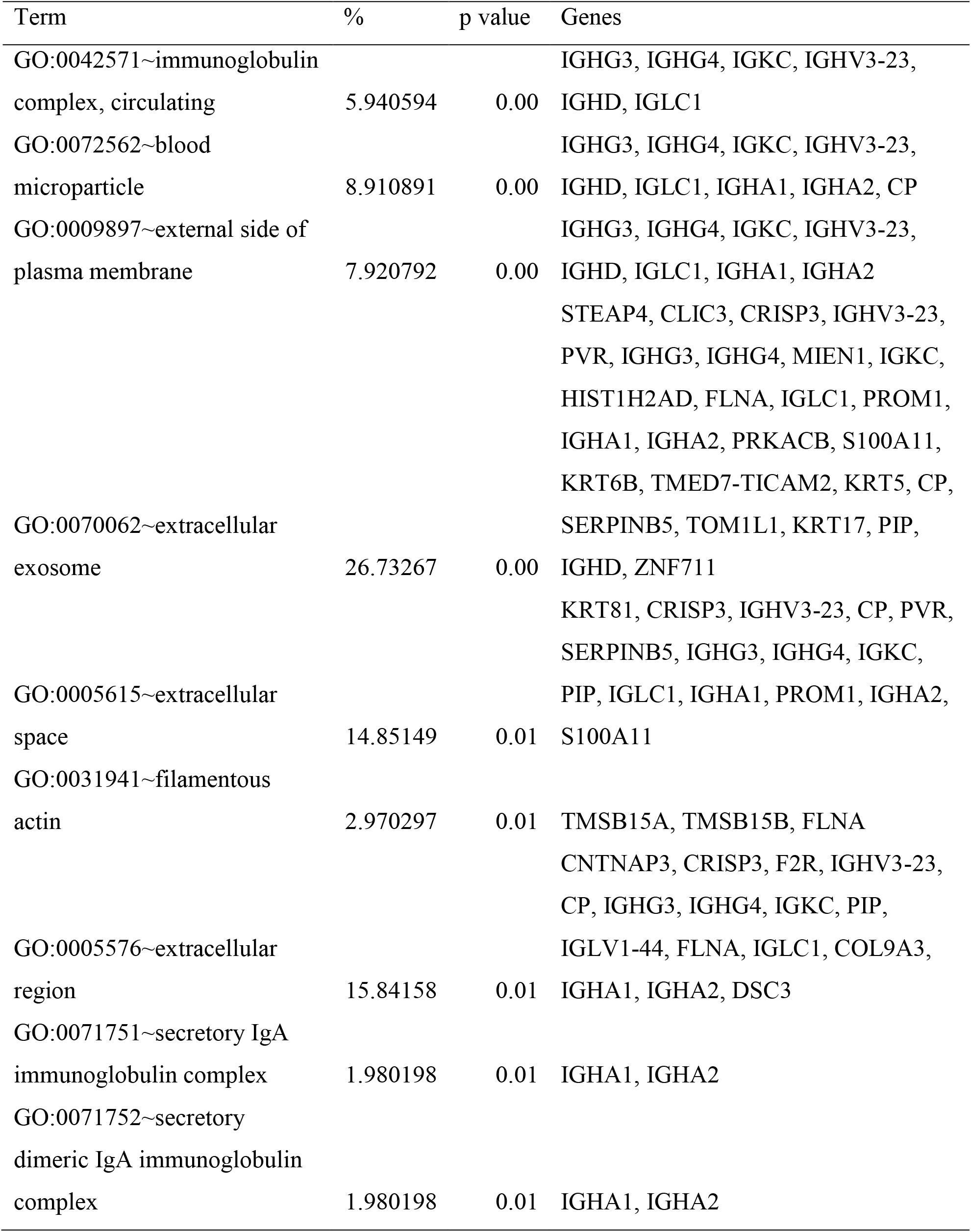
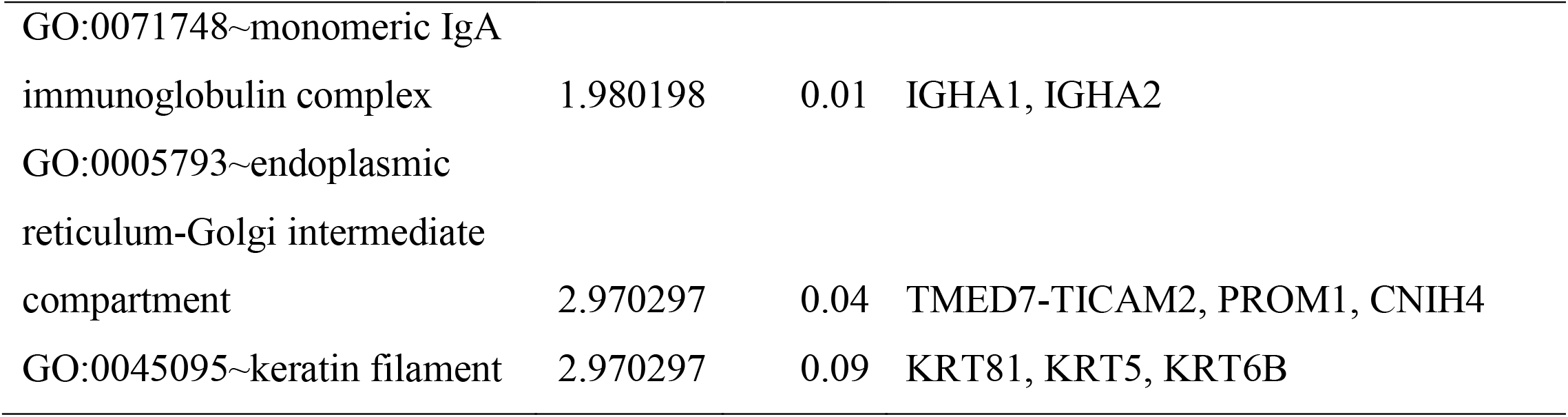
Gene Ontology analysis for the overlapping genes ∼ Cellular Components

**Table 5:**
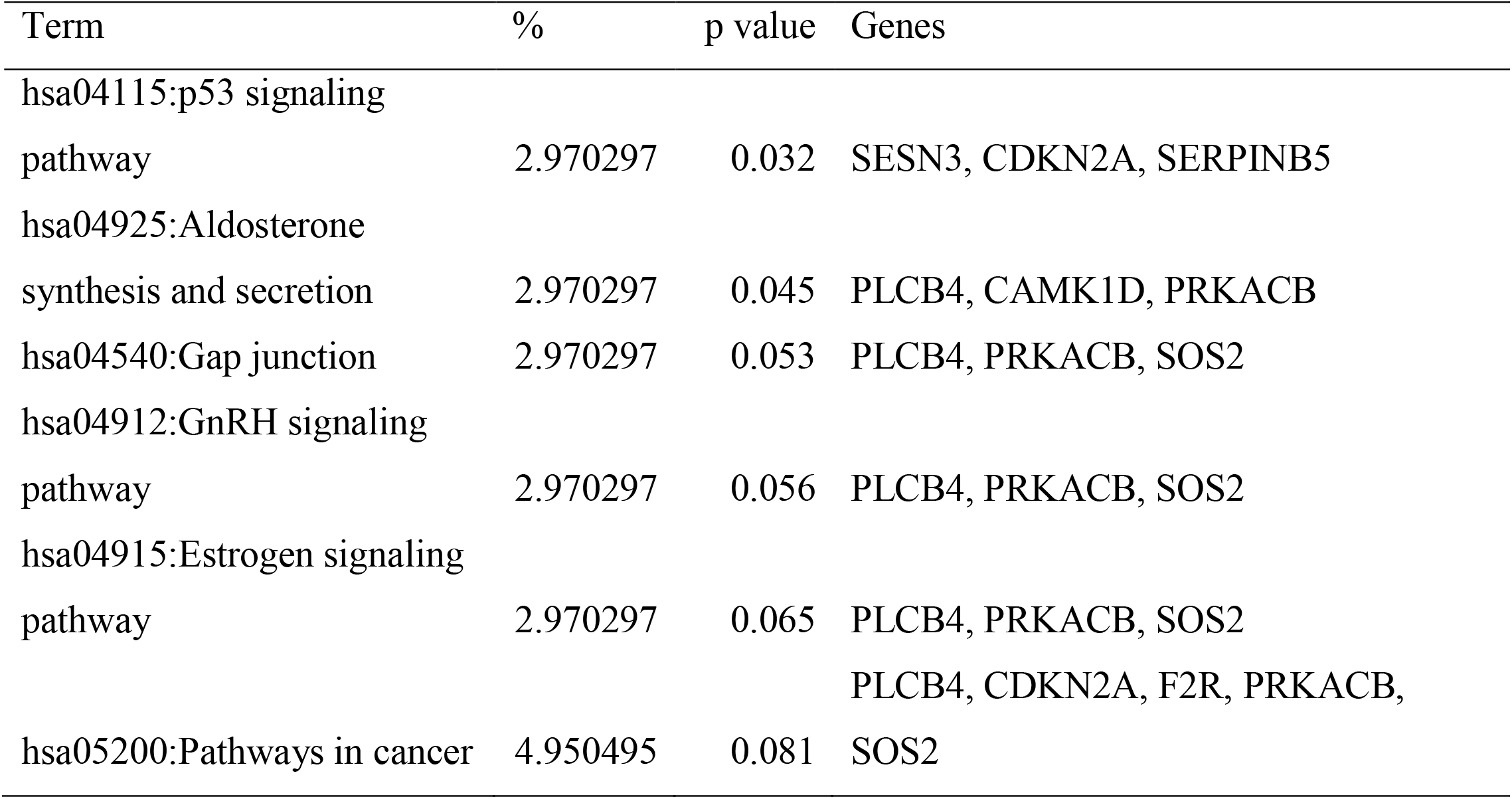
KEGG pathway analysis for the overlapping genes

**Figure 4a-d:**
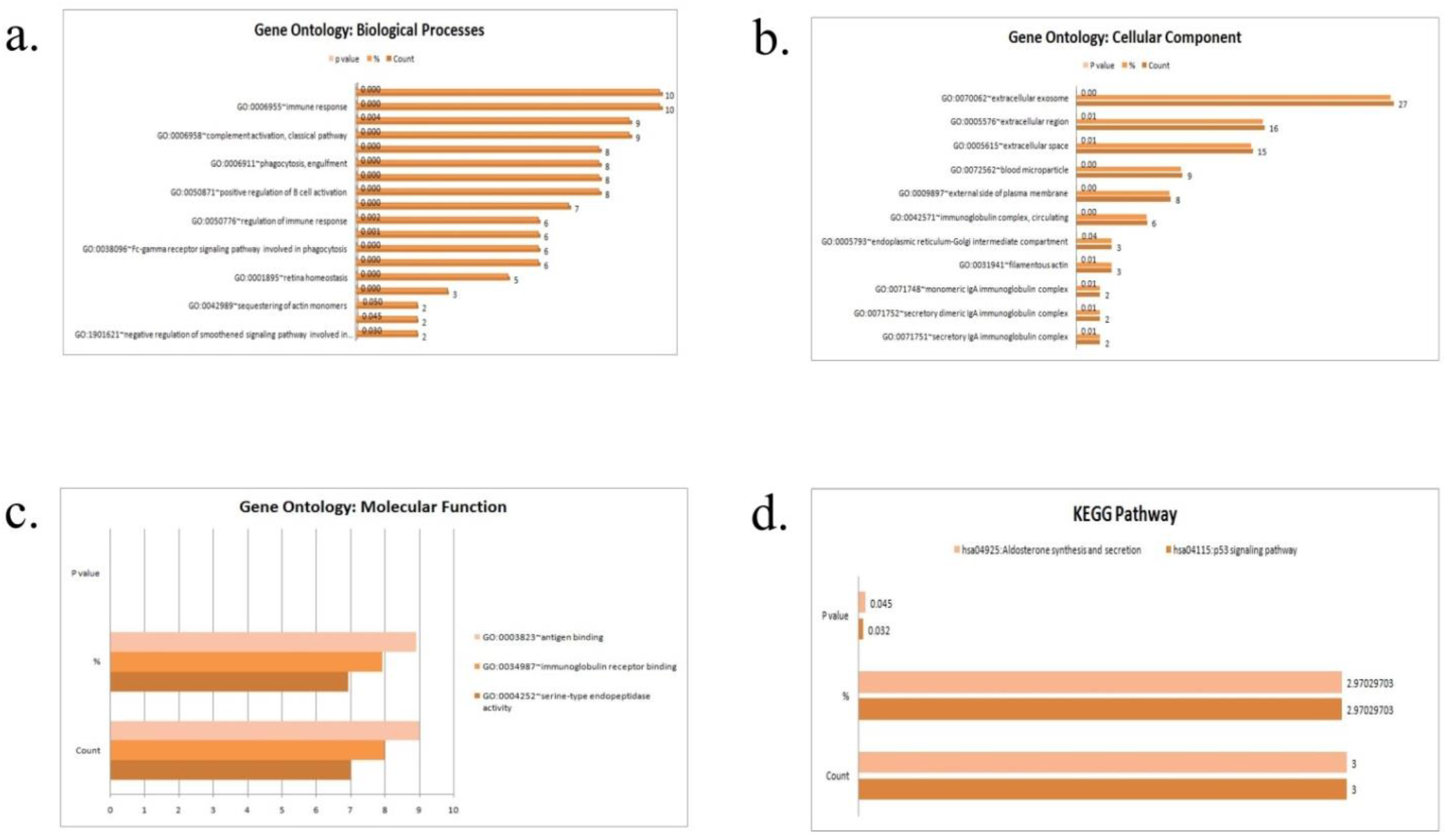
Gene Ontology (GO) and Kyoto Encyclopedia of Genes and Genomes (KEGG) pathway analysis by DAVID

### 3.3 PPI network construction and module analysis

To identify the interactions between the overlapping DEGs, online database STRING was used. A total of 86 DEGs as network nodes and 244 edges were used to construct and visualize the PPI network in Cytoscape (Figure 5).

**Figure 5:**
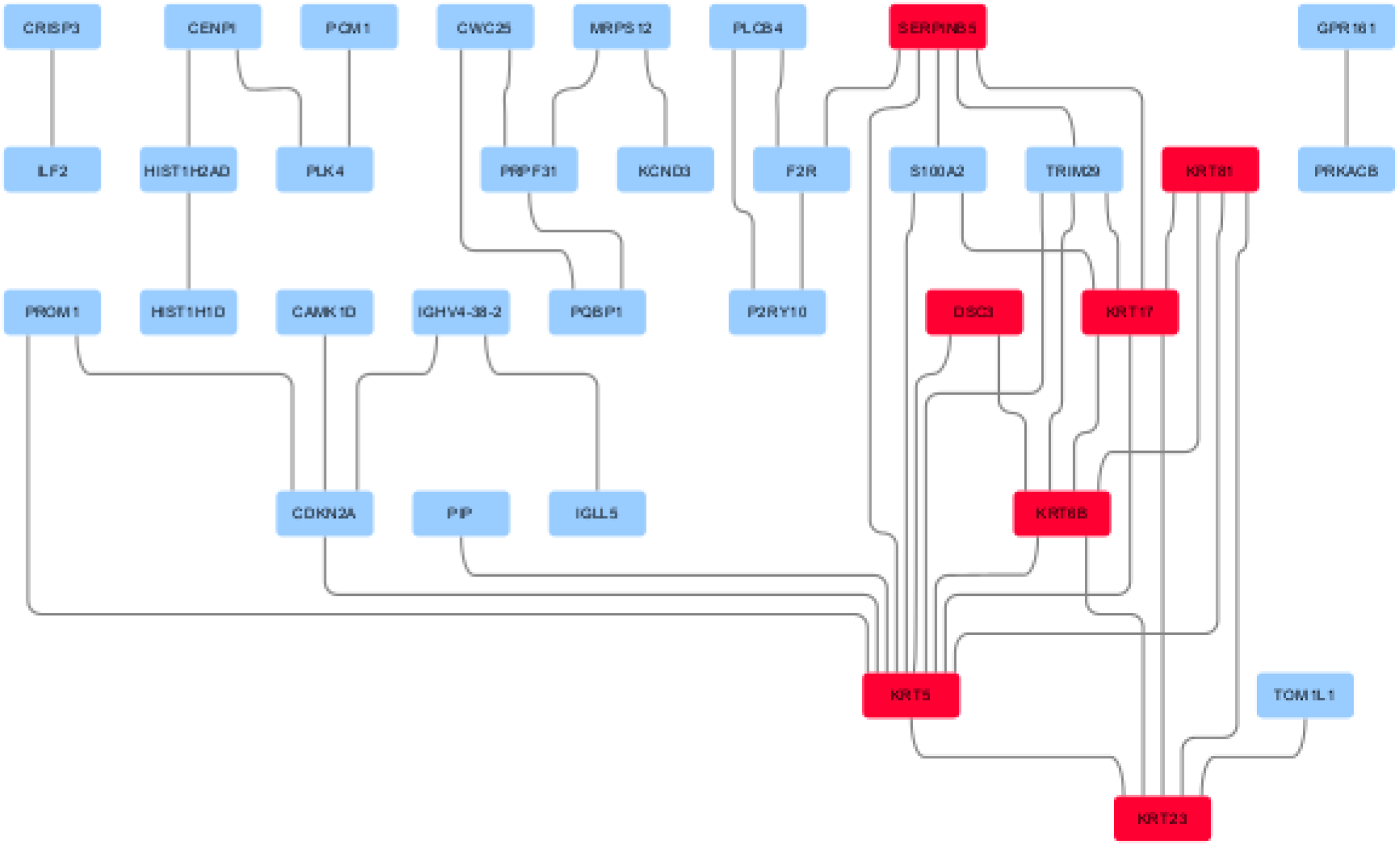
The protein-protein interaction (PPI) network for the selected hub genes visualized on Cytoscape

### 3.4 Hub gene analysis

Hub genes were identified using CytoHubba application, in which overlapping top 10 genes observed using twelve topological analysis methods, revealed that there were 21 hub genes, among which 13 genes were identified by at least three different methods as the candidate hub genes (Table 6).

**Table 6:**
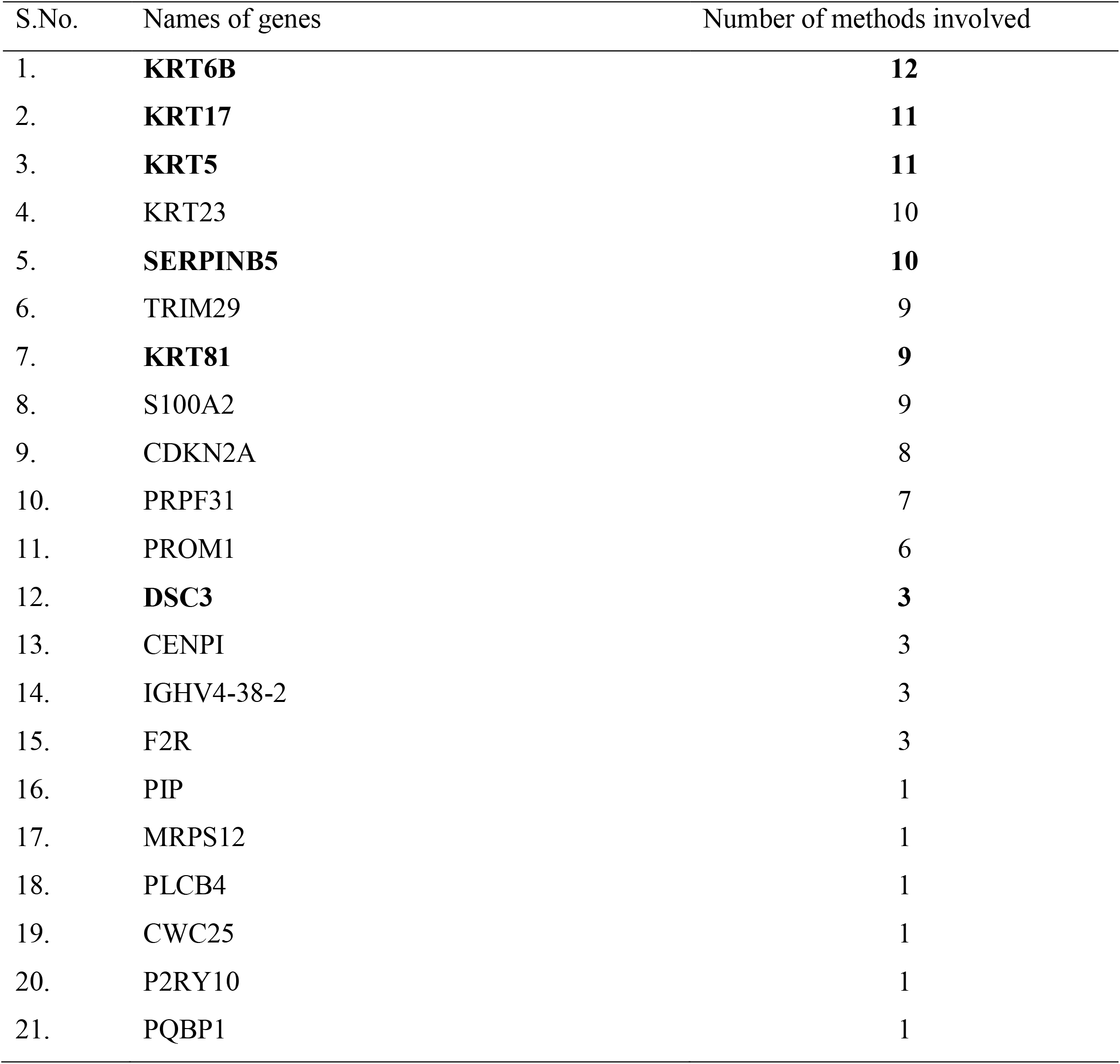
Hub gene analysis using CytoHubba plug-in to identify the key candidate genes

| S.No. | Names of genes | Number of methods involved |
| --- | --- | --- |
| 1. | **KRT6B** | **12** |
| 2. | **KRT17** | **11** |
| 3. | **KRT5** | **11** |
| 4. | KRT23 | 10 |
| 5. | **SERPINB5** | **10** |
| 6. | TRIM29 | 9 |
| 7. | **KRT81** | **9** |
| 8. | S100A2 | 9 |
| 9. | CDKN2A | 8 |
| 10. | PRPF31 | 7 |
| 11. | PROM1 | 6 |
| 12. | **DSC3** | **3** |
| 13. | CENPI | 3 |
| 14. | IGHV4-38-2 | 3 |
| 15. | F2R | 3 |
| 16. | PIP | 1 |
| 17. | MRPS12 | 1 |
| 18. | PLCB4 | 1 |
| 19. | CWC25 | 1 |
| 20. | P2RY10 | 1 |
| 21. | PQBP1 | 1 |

**Table 7:** Common miRNA targeting hub genes

| S.No. | Gene | Targeting miRNA |
| --- | --- | --- |
| 1. | DSC3 KRT17 KRT5 KRT6B<br>SERPINB5 | hsa-mir-155-5p |
| 2. | DSC3 KRT17 SERPINB5 | hsa-mir-103a-3p |
| 3. | DSC3 KRT6B SERPINB5 | hsa-mir-16-5p<br>hsa-let-7b-5p |
| 4. | KRT17 KRT5 SERPINB5 | hsa-mir-1343-3p |
| 5. | KRT17 KRT81 SERPINB5 | hsa-mir-1-3p<br>hsa-mir-194-5p |
| 6. | DSC3 KRT17 | hsa-mir-124-3p |
| 7. | DSC3 KRT81 | hsa-mir-129-2-3p<br>hsa-mir-522-5p<br>hsa-mir-27a-3p |
| 8. | DSC3 SERPINB5 | hsa-mir-107 |
| 9. | KRT17 KRT81 | hsa-mir-200b-3p |
| 10. | KRT17 SERPINB5 | hsa-mir-429 |
| 11. | KRT5 KRT81 | hsa-mir-146a-5p |
| 12. | KRT5 SERPINB5 | hsa-mir-335-5p<br>hsa-mir-21-5p |
| 13. | KRT6B SERPINB5 | hsa-mir-30a-5p |

### 3.5 Survival analysis

Association between hub gene expressions with OSof BC patients were analysed to examine the role of hub genes in BC prognosis by grouping the population into high expression and low expression categories, six genes showed a significant association between differential expressions with shorter OS among patients with BC. These six hub genes-DCS3 (HR=0.72, p=0.045), KRT17 (HR=0.51, p=0.002), KRT5 (HR=0.71, p=0.035), KRT6B (HR=0.57, p<0.001), KRT81 (HR=0.70, p=0.029) and SERPINB5 (HR=0.68, p=0.017) may serve as prognostic biomarkers to determine the severity of BC patients (Figure 6a-f). The expressions of the six hub genes in BC tissue compared with normal tissue are shown in Figure 7a-f.

**Figure 6a-f:**
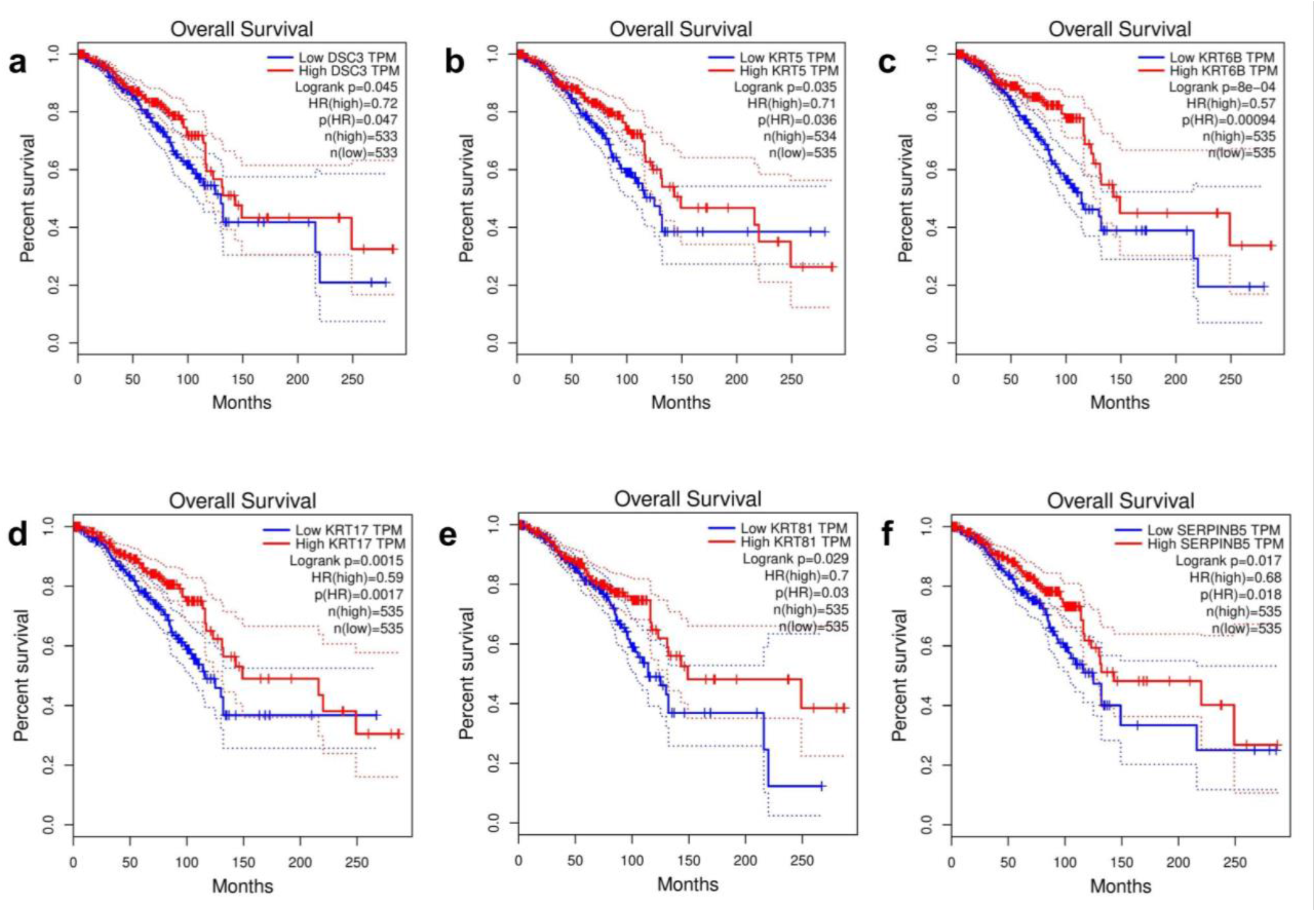
Associations of the six candidate hub genes with overall survival in breast cancer. Log rank *P*<0.05 is considered as statistically significant

**Figure 7a-f:**
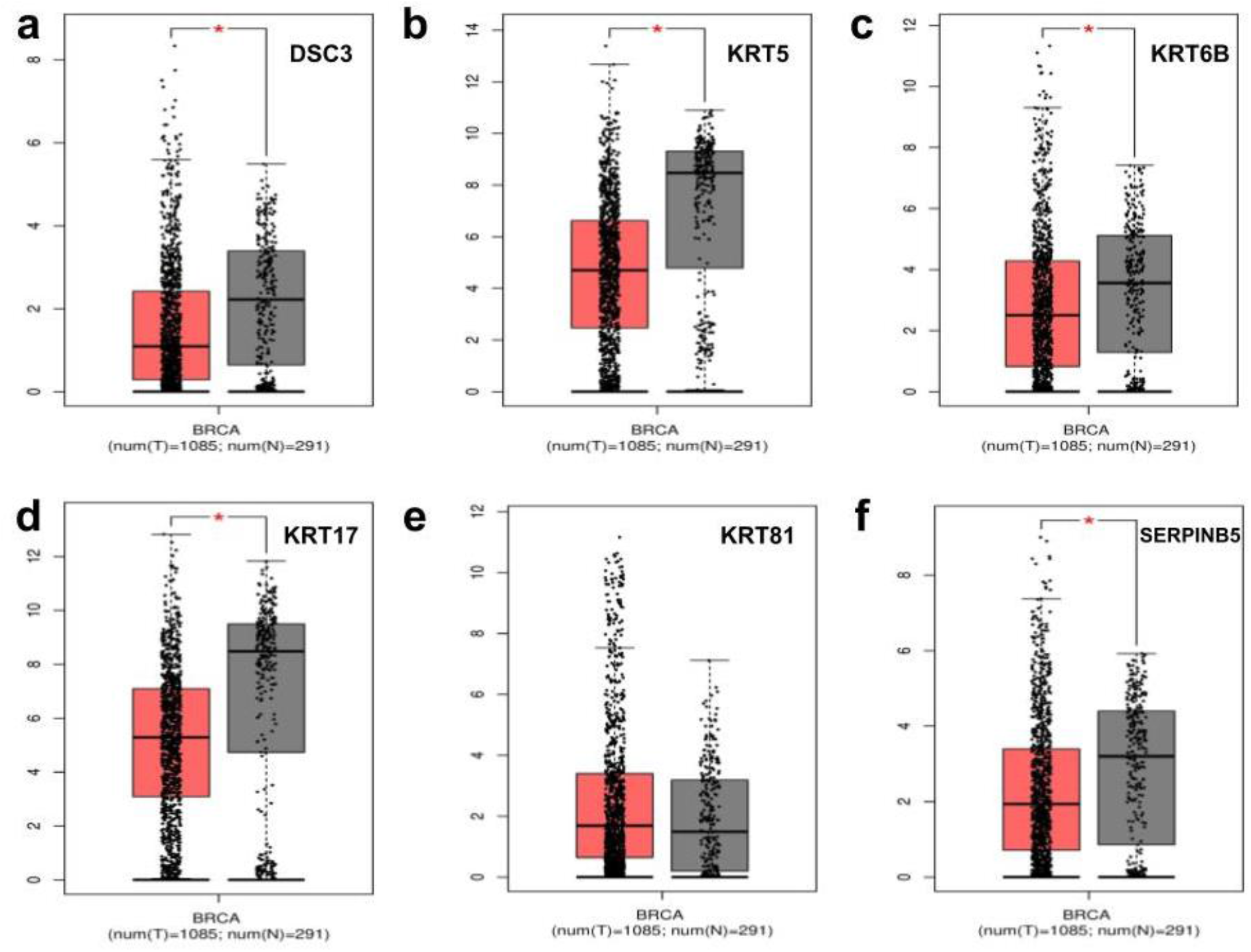
Differential expressions of the six candidate hub genes in breast cancer.

### 3.6 Hub genes targeting transcription factors and miRNA analysis

Hub genes targeting miRNA were predicted using miRNet, based on the correlation analysis between the hub genes and miRNAs. In the network, there were 6 genes and 83 miRNA, with 6 nodes and 110 edges. Out of these 83 miRNAs, hsa-miR-155-5p targetted all hub genes except KRT81. Further, using five different databases, gene-transcription factor network was analyzed, as depicted in Figure 8.

**Figure 8:**
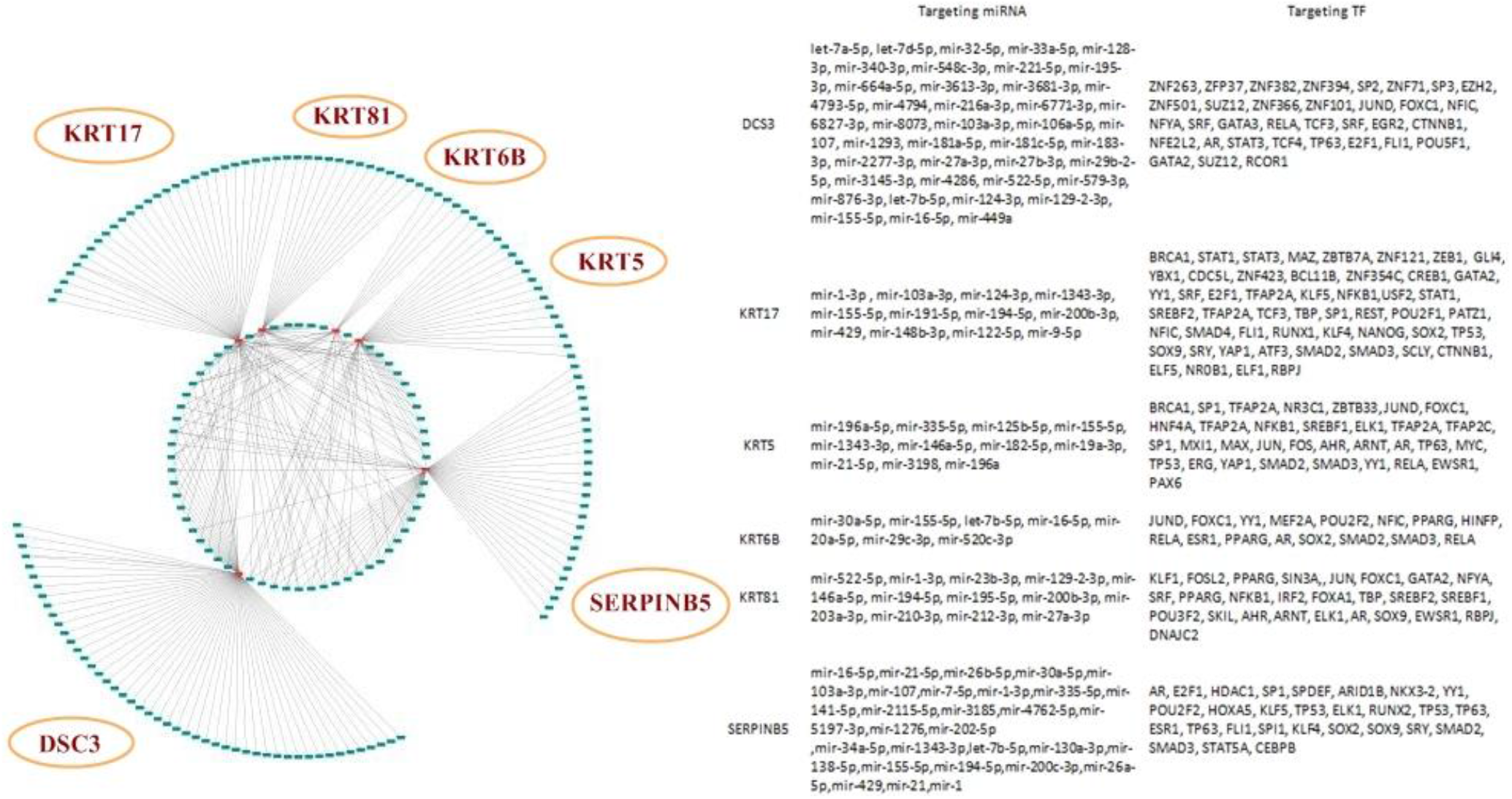
The hub gene-miRNA-transcription factor regulatory network

## 4. Discussion

The current study aimed to gain insight into the change in gene expression in LNM in BC through analyzing publicly available datasets. For this, we have identified the overlapping DEGs responsible for LNM in BC. Among these DEGs, six candidate hub genes, namely DSC3, KRT17, KRT5, KRT6B, KRT81 and SERPINB5, showed significant associations with poor OS in BC patients. Further, hub gene targeting miRNA were also identified. A total of 83 miRNA and 112 transcription factors were identified for these selected genes.

Axillary lymph node status is an important prognostic factor in BC. An increase in the number of metastatic lymph nodes involved is associated with a decrease OS as well as DFS [5,20]. Axillary lymph node status is also pivotal in determining the course of management e.g., radiotherapy in these patients [21,22]. Accordingly, the better prognosis of node-negative patients of BC is attributed to timely resection, before distant metastasis via the axillary lymphatics has occurred [23]. The numbers of these metastatic lymph nodes are those that are dissected by the surgeon and examined by the pathologist. However, various studies have shown that the metastatic lymph nodes are greater in number with an increasing number of removed nodes. Thus, it is difficult to assess the axillary lymph node status reliably without removing and identifying sufficient numbers of lymph nodes depending on the surgeon and pathologist [24-26]. Further, nodal status identification involves lymph node biopsy, which can potentially give false-negative results. Therefore, there is a need for an appropriate method to identify patients with and without LNM, which could also reduce the chances of comorbidity related to surgical evaluation. Identification of candidate genes related to nodal metastasis can aid in the better evaluation and subsequent management of these patients. Furthermore, BC metastasis has been known to be molecularly distinct from their primary tumor counterparts.

Desmocollin 3 (DSC3), a member of the cadherin superfamily of calcium-dependent cell adhesion molecules, is a desmosomal protein. DSC3 helps in maintaining tissue architecture; hence, their loss leads to a lack of adhesion and a gain of cellular mobility [27,28]. DSC3 is a p53 responsive gene, and its expression is down-regulated in BC cell lines and primary breast tumors, indicating that the loss of DSC3 expression is a common event in primary breast tumor specimens [27]. In esophageal adenocarcinoma (EACs) tissue samples and human EAC cell lines, a significant down-regulation (*P*<0.001) of the DSC3 mRNA levels has been observed. In addition, the EAC cell lines and tumor samples had aberrant promoter hypermethylation as compared to normal esophageal samples (*P*<0.001) [28]. DSC3 has also been implicated in LNM and cellular proliferation in oral squamous cell carcinoma (OSCC) through the regulation of β-catenin [29].

KRT genes are involved in the synthesis of keratin, which is a fibrous protein involved in the structure of epithelial cells. KRT5 is overexpressed in basal-like BC subtype tissue, as well as cell line and is associated with poor outcome [30]. It is also seen to be overexpressed in younger women with BC compared to older women [31] KRT6B, a key mediator of notch signalling [32], has been observed to be overexpressed in HCC and responsible for its progression. In BC, it is a favourable prognostic marker. Keratin 17 (KRT17) is a 48KDa type I intermediate filament, which is mainly expressed in epithelial basal cells. KRT17 is overexpressed in many malignant tumors and plays an important role in tumor development. It has been observed that KRT17 expression levels were significantly higher in lung cancer compared to normal lung tissues, and such a high expression of KRT17 predicted poor prognosis for patients with lung adenocarcinomas and was correlated with poor differentiation and lymphatic metastasis [33].

Overexpression of KRT17 enhanced, while its knockdown inhibited, the proliferation and invasiveness of lung cancer cells [33]. KRT17 promotes ESCC cell proliferation and migration, thus potentially contributing to metastasis. It also induces epithelial-mesenchymal transition through activation of AKT signalling [34]. Both KRT5 and KRT17 are positive markers for TNBCs [35]. KRT81 is found to be higher in BC tissue compared with normal mammary epithelial cells. In KRT81-knockdown MDA-MB231 cells, a decreased MMP9 activity was noticed, while along with decreased cell migration and invasion capabilities [36].

High immunoreactivity of SERPINB5 or Maspin (mammary serine protease inhibitor), is significantly linked to pre and post-CCRT advanced disease, lymphovascular invasion, and poor response to colorectal cancer (all *P*≤0.015). It is a metastasis suppressor gene. SERPINB5 overexpression is negatively associated with disease-specific survival (DSS), local recurrence-free survival (LRFS) and metastasis-free survival (MeFS) rates in colorectal cancer. It can also independently predict for DSS and MeFS in colorectal cancer (all *P*≤0.043) [37]. In fact, SERPINB5 is part of the metastatic epithelial-gene signature in BC and associated with an unfavourable prognosis [38]. In BC cell line, SERPINB5 inhibits tumour cell invasion and promotes cellular adhesion [39].

In this study, we also identified the miRNAs targeting these hub genes and hsa-miR155-5p was identified as targeting five hub genes. Similar findings have been reported in a clinical study involving 80 individuals, where BC patients showed a significant correlation with lymph node positive status and miR-155-5p expression [40]. Further, in-silico survival analysis found the genes to be significantly associated with OS, suggesting that these hub genes may serve as potential prognostic biomarkers and therapeutic targets for BC. However, there were certain limitations to our study. Firstly, when analyzing the DEGs, given the complexity of datasets in our study, it is difficult to consider demographic factors such as different age groups, ethnicity, geographical regions, as well as tumor staging and classification of all the patients. Secondly, according to the results, the six hub genes were all up or downregulated in BC, but the mechanism of their dysregulation is yet to be explored. Therefore, more pieces of evidences are required to find out their mechanistic foundation. The hsa-miR155-5p identified as common miR regulating the expression of hub genes needs further exploration in regards to its role in LNM. The transcription factors identified in this study could aid in the exploration of those pathways. Finally, this study has analyzed the expression levels and OS of the six hub genes with the help of bioinformatics, which may help to develop a biomarker panel for this patient population. However, to establish these hub genes as diagnostic or prognostic markers with high accuracy and specificity for BC, we need larger, prospective studies.

## Data Availability

Not applicable.

## List of abbreviations

BC: Breast Cancer
CCRT: Concurrent chemoradiotherapy
CHEA: Chip Enrichment Analysis
DAVID: database for annotation, visualization and integrated discovery
DEG: Differentially expressed genes
DFS: Disease-free survival
DSS: Disease specific survival
ENCODE: Encyclopedia of DNA elements
GEO: Gene Expression Omnibus
GEPIA: Gene Expression Profiling Interactive Analysis
GO: Gene Ontology
KEGG: Kyoto Encyclopedia of Genes and Genomes
LNM: Lymph Node Metastasis
LRFS: Local recurrence-free survival
Maspin: mammary serine protease inhibitor
MeFS: metastasis free survival
miRNA: MicroRNA
OS: Overall Survival
PPI: Protein-protein interaction
SLNB: sentinel lymph node biopsy
STRING: Search Tool for the Retrieval of Interacting Genes and proteins
TRRUST: Transcriptional Regulatory Relationships Unravelled by Sentence-based Text Mining

## Conflict of interest

The authors declare no conflicts of interest.

## Acknowledgement

The authors acknowledge the support of the Department of Biotechnology, Ministry of Science and Technology (no. DBT/2018/AIMS-J/994).

## Notes

### Competing Interest Statement

The authors have declared no competing interest.

### Funding Statement

No funding was received for this study.

### Author Declarations

Institutional Ethics Committee (IEC), All India Institute of Medical Sciences(AIIMS), Jodhpur

